# BUCAN: Bayesian Uncertainty-aware Classification with Attention Networks for Medical Images

**DOI:** 10.1101/2025.11.05.25339638

**Authors:** Abhinav Sagar

## Abstract

Accurate and reliable medical image classification is critical for clinical decision-making across diverse imaging modalities, including X-ray, CT, and MRI. Traditional convolutional neural networks often produce overconfident predictions, limiting their clinical trustworthiness. In this work, we propose an uncertainty-aware, attention-augmented neural network that integrates multi-scale SwirlAttention and FeedBackAttention modules with a Bayesian probabilistic classifier. This framework enables robust feature extraction, interpretable attention maps, and principled estimation of epistemic uncertainty. We evaluate our approach on four diverse datasets, including Diabetic Retinopathy, Kvasir, Skin Cancer, and fused multi-focal Oocyte images, covering a wide range of pathological and morphological variations. Extensive experiments demonstrate that our method outperforms state-of-the-art CNN and transformer-based baselines in terms of accuracy, calibration, and interpretability. Grad-CAM visualizations highlight clinically relevant regions, while uncertainty estimates provide actionable insights for ambiguous cases, making the framework suitable for reliable deployment in real-world clinical settings.

## Introduction

Medical image classification plays a critical role in modern clinical workflows by enabling automated diagnosis, early disease detection, and treatment planning across a wide range of imaging modalities. Despite significant advances with deep learning, conventional convolutional neural networks (CNNs) often operate under deterministic assumptions, producing overconfident predictions when confronted with noisy, ambiguous, or out-of-distribution medical images. This limitation is particularly concerning in safety-critical applications, where diagnostic errors can have severe clinical consequences.

Recent research has shown that incorporating attention mechanisms can improve both predictive accuracy and interpretability. Channel and spatial attention modules, such as SE (Squeeze-and-Excitation) (Hu, Shen, and Sun 2018) and CBAM (Convolutional Block Attention Module) (Woo et al. 2018), enable networks to adaptively emphasize informative feature maps and focus on the most relevant regions within an image. By jointly modeling inter-channel relationships and spatial dependencies, these mechanisms enhance the network’s ability to discriminate between visually similar classes and provide more interpretable feature representations for clinical decision-making. However, existing attention-based methods still face several challenges. Many architectures tend to overfit on small medical datasets due to their high model complexity, while others struggle to generalize across imaging modalities with varying contrast, resolution, and noise characteristics. Furthermore, most attention mechanisms provide deterministic outputs without quantifying uncertainty, limiting their reliability in critical diagnostic scenarios where confidence estimation is essential.

In this study, we propose a novel framework that integrates multi-scale attention with a Bayesian probabilistic classifier for robust medical image classification. Our approach is evaluated on four diverse datasets spanning multiple organs and imaging modalities: Diabetic Retinopathy fundus images, Kvasir endoscopic images, Skin Cancer dermoscopic images, and a fused multi-focal Oocyte dataset. Extensive experiments demonstrate that our method not only achieves state-of-the-art classification performance but also provides interpretable attention maps and meaningful uncertainty estimates, making it particularly suitable for clinical deployment.

To further explore the interpretability of our framework, we visualize the Grad-CAM-based attention maps alongside uncertainty quantification for oocyte images, as illustrated in Fig. 1. The highlighted regions in the attention maps represent discriminative features that significantly contribute to the model’s decision-making process. For oocyte classification, the model primarily focuses on morphological structures and texture patterns that indicate variations in oocyte quality.

**Figure 1:**
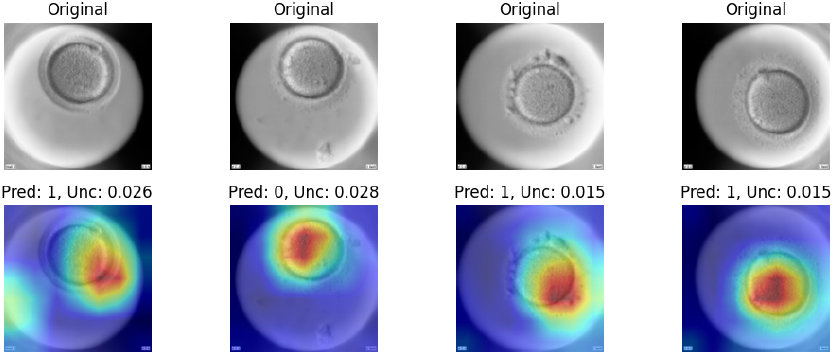
Illustration of GradCAM-based attention maps along with predicted labels and uncertainty quantification using the Oocytes dataset.

The main contributions of this work are summarized as follows:

1. We introduce a novel attention-augmented residual network that integrates SwirlAttention and FeedBackAttention modules to capture multi-scale spatial and channel-wise dependencies in medical images.
2. We incorporate a Bayesian probabilistic classifier using Bayes-by-Backprop, enabling reliable epistemic uncertainty estimation alongside discriminative predictions.
3. We conduct comprehensive experiments on four diverse medical imaging datasets, demonstrating improved accuracy, calibration, and interpretability compared to state-of-the-art CNN and transformer-based baselines.
4. We provide qualitative insights using Grad-CAM visualizations, showing that the network consistently attends to clinically relevant regions and highlighting cases where predictions may be less reliable.

Overall, this work presents a unified framework for robust, interpretable, and uncertainty-aware medical image classification, addressing key limitations of conventional deterministic approaches while providing practical tools for real-world clinical applications.

### Related Work

Medical image classification has witnessed rapid advancements with the adoption of deep learning, ranging from convolutional neural networks (CNNs) to vision transformers (ViTs) and hybrid architectures. Early works focused on CNNs for various modalities, demonstrating strong performance in tasks such as retinal fundus analysis, dermoscopic lesion classification, and brain tumor detection (Tajbakhsh et al. 2016; Swati et al. 2019). Residual and densely connected architectures further improved feature representation and gradient flow, with models such as ResGANet (Cheng et al. 2022) and DenseNet (Huang et al. 2017) achieving state-of-the-art results on multiple datasets.

Recent studies have emphasized attention mechanisms to enhance feature discriminability and interpretability. Lesion-aware networks (Fang et al. 2019), multi-scale pyramid fusion models (Wen et al. 2025), and hierarchical attentive fusion approaches (Abdar et al. 2022) have shown that guiding the network to focus on clinically relevant regions improves both accuracy and reliability. Concurrently, uncertainty-aware methods, including BARF (Abdar et al. 2021) and BayTTA (Sherkatghanad et al. 2025), have highlighted the importance of estimating epistemic and aleatoric uncertainty for robust clinical deployment.

Transformers and hybrid CNN-Transformer architectures have also gained traction in medical imaging due to their capacity for capturing long-range dependencies. Models such as MedViT (Manzari et al. 2023), MedMamba (Yue and Li 2024), TransMed (Dai, Gao, and Liu 2021), and MedTransNet (Shaik et al. 2024) leverage self-attention for global context modeling, while hybrid designs like EFFResNet-ViT (Hussain et al. 2025) and DBCvT (Li, Feng, and Xia 2024) combine convolutional feature extraction with transformer-based token modeling for improved performance on multi-modal datasets.

Several works have focused on model calibration and reliable uncertainty estimation to mitigate overconfident predictions, particularly in class-imbalanced scenarios (Rajaraman, Ganesan, and Antani 2022; Liang et al. 2020; Ju et al. 2022). Dynamic fusion networks such as DAFNet (Cai et al. 2025) and multi-task Mamba variants (Wu and Gou 2025) further enhance robustness by adaptively integrating multi-scale and multi-modal information.

Despite these advances, challenges remain in simultaneously achieving high classification accuracy, interpretable attention, and reliable uncertainty estimation across diverse medical imaging modalities. The proposed framework addresses these gaps by integrating multi-scale attention modules with a Bayesian probabilistic classifier, offering both strong discriminative capability and trustworthy uncertainty quantification. Our method is evaluated across four distinct datasets, including fundus, dermatology, endoscopy, and fused multi-focal microscopy images, demonstrating broad applicability and improved reliability compared to existing approaches.

## Methodology

### Problem Definition

Medical image classification aims to automatically categorize medical scans or images into clinically meaningful classes, such as disease presence, tissue type, or anatomical region, based on their visual and structural characteristics. Formally, let the dataset be defined as

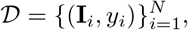

where **I**_*i*_ ∈ *R*^3×*H*×*W*^ or **I**_*i*_ ∈ *R*^1×*H*×*W*^ represents a medical image (e.g., MRI, CT, or histopathology image) and *y*_*i*_ ∈ *Y* = {1, 2, …, *C*} denotes its ground-truth diagnostic label among *C* possible disease categories. The goal of the model is to learn a mapping function.

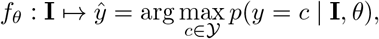

parameterized by *θ*, that accurately predicts the correct class label ŷ for each input image.

Traditional CNNs achieve high discriminative power but operate under deterministic weight assumptions, which often yield overconfident predictions in the presence of noisy or out-of-distribution medical data. This limitation is particularly critical in clinical settings where predictive confidence directly affects diagnostic reliability and downstream decision-making.

To mitigate this issue, we introduce a Bayesian uncertainty-aware framework that models the weight parameters *w* of the classification layer as probability distributions rather than fixed values. The predictive distribution for a new unseen image **I**^∗^ is expressed as:

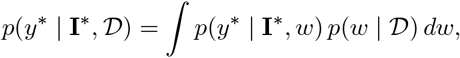

Where *p*(*w* | *D*) represents the posterior over the weights given the training data. Since this integral is intractable, it is approximated through a variational posterior *q*(*w θ*) using the Bayes-by-Backprop principle.

Thus, the learning objective is to infer both discriminative parameters and uncertainty estimates by minimizing the negative evidence lower bound (ELBO):

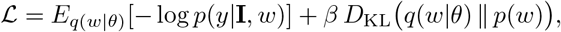

Where the first term corresponds to the expected classification loss and the second term regularizes the approximate posterior toward the prior distribution.

In summary, the problem of medical image classification is reformulated as learning a robust, uncertainty-aware mapping.

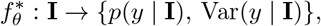

That outputs both diagnostic probabilities and corresponding uncertainty estimates. This design ensures reliable and interpretable predictions, a crucial property for safety-critical medical decision-making.

### Network Architecture

The proposed architecture integrates deterministic and probabilistic reasoning through a hierarchical attention-based residual backbone coupled with a Bayesian classification head. The overall design aims to achieve robust feature representation under uncertainty while maintaining efficient spatial–channel interactions.

#### 1) Swirl Attention

To enhance spatial contextual reasoning, we introduce the **Swirl Attention** module, which performs multi-scale depthwise convolutions with kernel sizes {3, 5, 7, 9}. These convolutions emulate a “swirling” expansion of the receptive field, enabling the model to capture both fine and coarse spatial dependencies. The outputs from each scale are concatenated and fused through a 1 *×* 1 convolution followed by batch normalization:

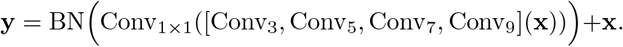

This residual formulation ensures stable feature propagation while enriching contextual awareness.

#### 2) FeedBack Attention

We propose a novel **FeedBack Attention** mechanism to jointly refine spatial and channel-wise activations. It consists of (i) a channel attention path based on global average pooling and two-layer bottleneck transformation, and (ii) a spatial attention path computed from the concatenation of average- and max-pooled channel responses. Unlike conventional dual-branch attention (e.g., CBAM (Woo et al. 2018)), the FeedBack Attention reuses the spatially modulated features to re-estimate the channel weights, thus forming a feedback loop that progressively refines salient feature activations. The joint attention is applied as:

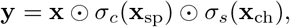

Where *σ*_*c*_ and *σ*_*s*_ denote channel and spatial attention maps, respectively.

#### 3) Residual Attention Blocks

Each residual unit integrates both the Swirl and FeedBack attention modules, providing adaptive feature recalibration and multi-scale context aggregation. When downsampling is required, a 1 × 1 convolutional skip connection is employed to match dimensions. The block formulation is given by:

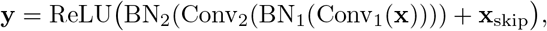

Where intermediate activations are modulated by the two attention branches.

#### 4) Bayesian Classification Head

To quantify epistemic uncertainty, the deterministic backbone is followed by a Bayesian inference layer using the Bayes-by-Backprop framework. The final fully connected layer is replaced by a **BayesLinear** module, where each weight and bias is modeled as a Gaussian distribution parameterized by (*µ, ρ*), with *σ* = log(1 + exp(*ρ*)) ensuring positivity. During training, weights are sampled as:

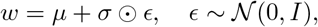

And optimized via the evidence lower bound (ELBO), incorporating a Kullback–Leibler divergence term against a standard normal prior. This probabilistic head enables calibrated uncertainty estimation at inference.

#### 5) Overall Architecture

The complete model begins with a convolutional stem followed by four hierarchical Residual Attention Blocks with progressive channel expansion (32 → 64 → 128 → 256). A global average pooling layer and dropout regularization precede the Bayesian classifier. The integrated attention hierarchy and probabilistic inference enable robust and interpretable decision-making under data noise and domain variability.

The complete network architecture diagram is shown in Figure 2.

**Figure 2:**
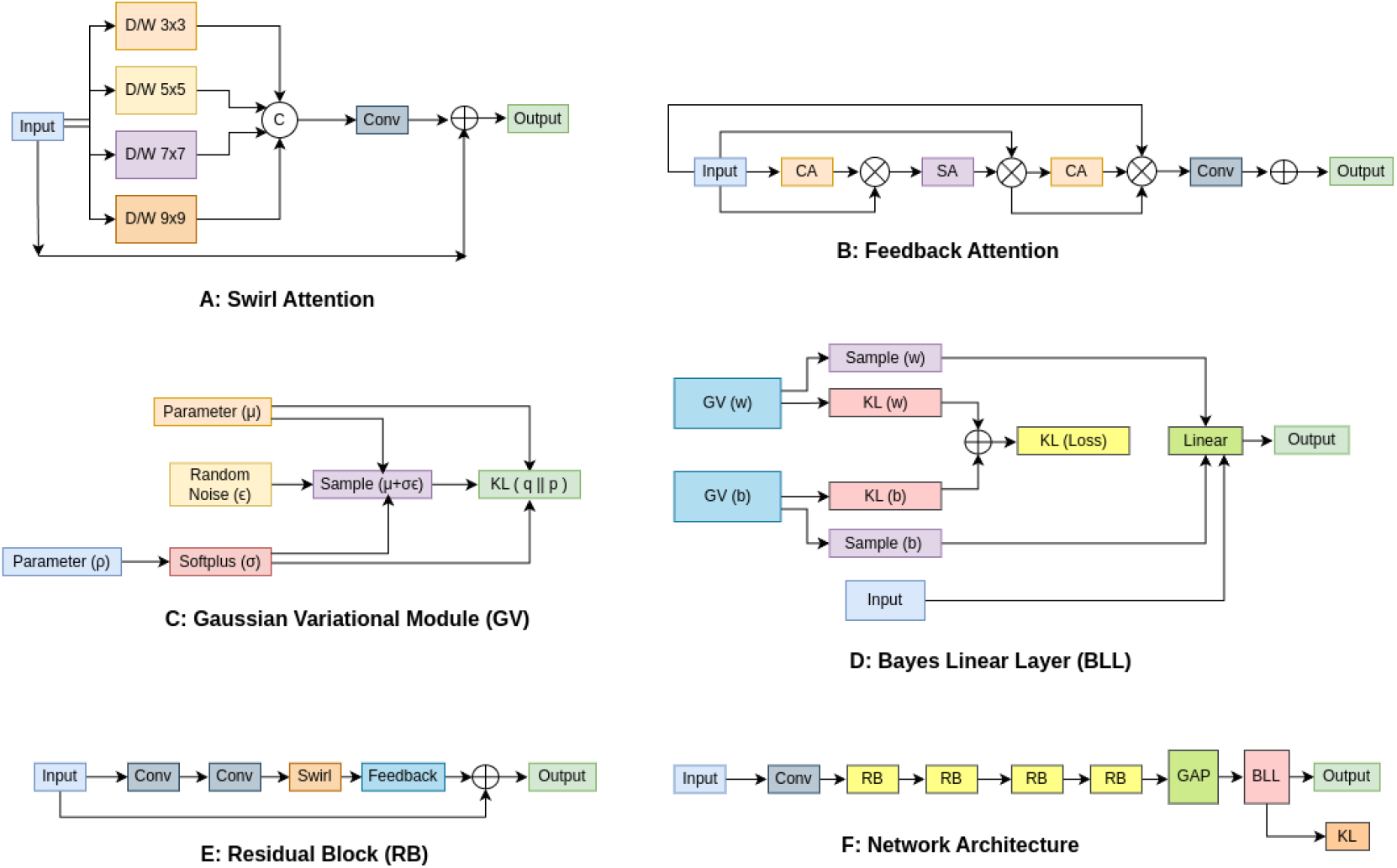
Illustration of the proposed network architecture and its components - A: Swirl Attention. B: Feedback Attention. C: Gaussian Variational Module. D: Bayes Linear Layer. E: Residual Block. F: Complete Network Architecture. Here, D/W, Conv, CA, SA, KL, Linear, and GAP denote Depthwise Convolution, Regular Convolution, Channel Attention, Spatial Attention, KL Divergence, Fully-Connected Layer, and Global-Average-Pooling Layer, respectively.

### Loss Function

The proposed framework employs a Bayesian variational learning strategy to jointly optimize deterministic feature representations and probabilistic model parameters. Given the Bayes-by-Backprop formulation of the final classification layer, each weight and bias is represented by a variational distribution parameterized by its mean *µ* and log-scale parameter *ρ*. This allows the model to capture epistemic uncertainty through stochastic sampling during training.

Formally, let *q*(*w* | *θ*) denote the approximate posterior distribution over the network parameters *w*, where *θ* = {*µ, ρ*}, and let *p*(*w*) represent a standard normal prior. The learning objective is derived from the maximization of the evidence lower bound (ELBO), which can be equivalently expressed as a minimization problem:

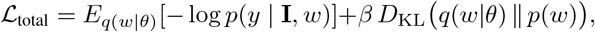

where: - The first term, *E*_*q*(*w* | *θ*)_[−log *p*(*y* | **I**, *w*)], corresponds to the expected negative log-likelihood of the correct class and serves as the **classification loss**. - The second term, *D*_KL_(*q*(*w* | *θ*) ∥ *p*(*w*)), represents the **Kull-back–Leibler (KL) divergence** between the approximate posterior and the prior, acting as a regularizer that penalizes overconfident posterior distributions. - The scalar *β* balances predictive accuracy and uncertainty regularization; empirically, *β* ∈ [10^−4^, 10^−2^] yields stable convergence.

For medical image classification, the likelihood term is modeled using the categorical cross-entropy loss:

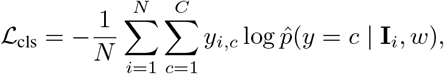

where *y*_*i,c*_ denotes the one-hot encoded ground-truth label and 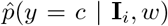 is the predicted class probability. The final loss function can thus be written as:

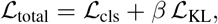

with

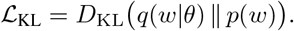

During training, the reparameterization trick

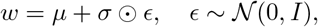

It is used to enable gradient-based optimization through stochastic sampling. The model parameters *θ* are updated via backpropagation to minimize ℒ_total_.

This combined objective ensures that the model simultaneously learns discriminative features and captures epistemic uncertainty. As a result, the proposed loss formulation encourages reliable predictions with calibrated confidence—an essential property for robust decision-making in medical image analysis.

## Experiments

### Dataset

To comprehensively evaluate the effectiveness and generalizability of the proposed medical image classification framework, four publicly available and custom-prepared datasets were utilized. These datasets span multiple imaging modalities, organs, and diagnostic tasks, ensuring robust validation across diverse clinical applications.

1. **Diabetic Retinopathy (DR):** The Diabetic Retinopathy dataset contains high-resolution retinal fundus images annotated with five severity levels: *No DR, Mild, Moderate, Severe*, and *Proliferative DR*. Specifically, the dataset includes 1,805 images without DR, 370 mild cases, 999 moderate cases, 193 severe cases, and 295 proliferative DR cases (Asia Pacific Tele-Ophthalmology Society (APTOS) 2019).
2. **Kvasir:** The Kvasir dataset consists of endoscopic gastrointestinal (GI) tract images annotated for both anatomical landmarks and pathological findings. It contains 1,000 images per category across eight classes: dyed-lifted polyps, dyed-resection margins, esophagitis, normal cecum, normal pylorus, normal z-line, polyps, and ulcerative colitis (Pogorelov et al. 2017).
3. **Skin Cancer (HAM10000):** The HAM10000 dataset comprises dermoscopic images of skin lesions spanning nine diagnostic categories, covering both benign and malignant conditions. Specifically, it includes 130 images of actinic keratosis, 392 basal cell carcinoma, 111 dermatofibroma, 454 melanoma, 373 nevus, 478 pigmented benign keratosis, 80 seborrheic keratosis, 197 squamous cell carcinoma, and 142 vascular lesions (Codella et al. 2019).
4. **Oocyte Multi-Focal Fusion Dataset:** A specialized private oocyte imaging dataset was constructed by fusing **11 focal plane images** into a single high-information composite using an adaptive fusion strategy. The dataset is categorized into four biologically meaningful classes representing distinct oocyte maturation stages.

### Implementation Details

#### Dataset and Preprocessing

All images were resized to 224 × 224 pixels and normalized using standard ImageNet statistics. For cross-validation experiments, a 5-fold stratified splitting strategy was employed to preserve class distributions across folds.

#### Training

Models were optimized using the Adam optimizer with an initial learning rate of 1 × 10^−4^. Training was conducted for 100 epochs per fold in the cross-validation setting using a batch size of 4. During inference, Monte Carlo Dropout with 30 stochastic forward passes was utilized to estimate epistemic uncertainty.

#### Software and Hardware

The framework was implemented in Python 3.10 using PyTorch 2.1, torchvision, and scikit-learn. All training and evaluation were performed on a workstation equipped with an NVIDIA A100 GPU (CUDA 11.8), 32 GB RAM, and an Intel Xeon CPU.

#### Evaluation Metrics

Model performance was assessed using accuracy, precision, recall, F1-score, Matthews correlation coefficient (MCC), and ROC-AUC. Calibration quality was evaluated using the Brier score, Expected Calibration Error (ECE), and reliability diagrams. Confusion matrices, ROC curves, and precision-recall curves were generated for qualitative assessment. Additionally, Grad-CAM was applied to visualize attention maps and interpret model decision-making.

### Comparison Approaches

To evaluate the effectiveness of the proposed uncertainty-aware, attention-augmented framework, we benchmark it against several state-of-the-art models for medical image classification. The selected baselines encompass a broad range of architectures, including conventional convolutional neural networks (CNNs), lightweight mobile networks, and modern transformer-based approaches, providing a comprehensive performance comparison.

#### Quantitative Performance

Quantitative evaluation was conducted on four diverse medical imaging datasets: Diabetic Retinopathy (Asia Pacific Tele-Ophthalmology Society (APTOS) 2019), Kvasir (Pogorelov et al. 2017), Skin Cancer (HAM10000) (Codella et al. 2019), and the private Oocyte Multi-Focal Fusion dataset. The comparison approaches include the following representative models:

- **CNN-based models:** DenseNet121 (Huang et al. 2017), ResNet50 (He et al. 2016), ResNeXt101 (Xie et al. 2017), RepVGG (Ding et al. 2021), ShuffleNetV2 (Ma et al. 2018), MobileNetV3 (Howard et al. 2019), EfficientNetB0 (Tan and Le 2019), and InceptionNextTiny (Yu et al. 2024).
- **Transformer-based and hybrid models:** SimpleViT (Beyer, Zhai, and Kolesnikov 2022), RegionViT (Chen, Panda, and Fan 2021), VAN (Guo et al. 2023), EfficientFormerL1 (Li et al. 2022), MobileViTXXS (Mehta and Rastegari 2021), MLPMixer (Tolstikhin et al. 2021), and ConvMixer (Trockman and Kolter 2022).

All models were trained and evaluated under the same preprocessing, data augmentation, and cross-validation protocols, ensuring a fair comparison.

#### Qualitative Performance

Figure 3 offer qualitative insights into the model’s performance. Figure 3 A depicts the confusion matrix using the Diabetic-Retinopathy dataset, showing that the classifier achieves high true positive rates across most classes with only a few misclassifications. Figure 3 B illustrates the ROC–AUC curves using the skin cancer dataset, indicating strong discriminative power between classes, as most curves approach the ideal top-left region. Th right diagram in Figure 3 C presents the precision–recall curves using the Kvasir dataset, demonstrating that the model sustains high precision even at elevated recall levels.

**Figure 3:**
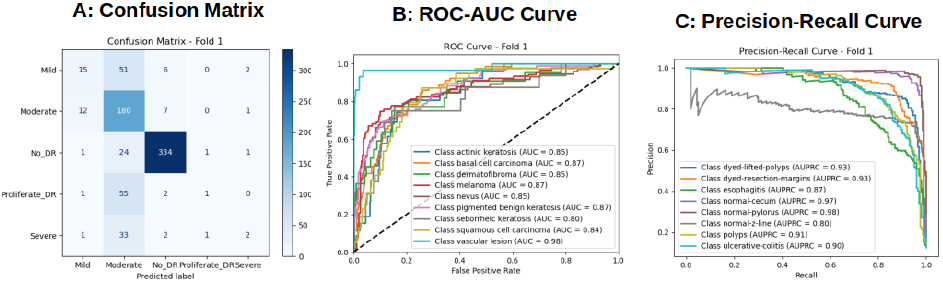
From left to right - A: Confusion matrix using the diabetic retinopathy dataset, B: ROC-AUC curve using the skin cancer dataset, and C: Precision-Recall curve using the Kvasir dataset

### Attention Visualization

To provide insights into the decision-making process of the proposed model, Grad-CAM (Selvaraju et al. 2017) was employed to generate visual explanations of class-specific activations. For each input image, heatmaps were overlaid on the original images to highlight regions that contributed most strongly to the predicted class.

The Grad-CAM visualizations reveal that the model consistently attends to clinically relevant regions. For example, in retinal images, the model highlights microaneurysms, hemorrhages, and exudates corresponding to diabetic retinopathy severity as in Figure 4. Similarly, in dermoscopic images, attention is concentrated on lesion boundaries and abnormal tissue structures as in Figure 5.

**Figure 4:**
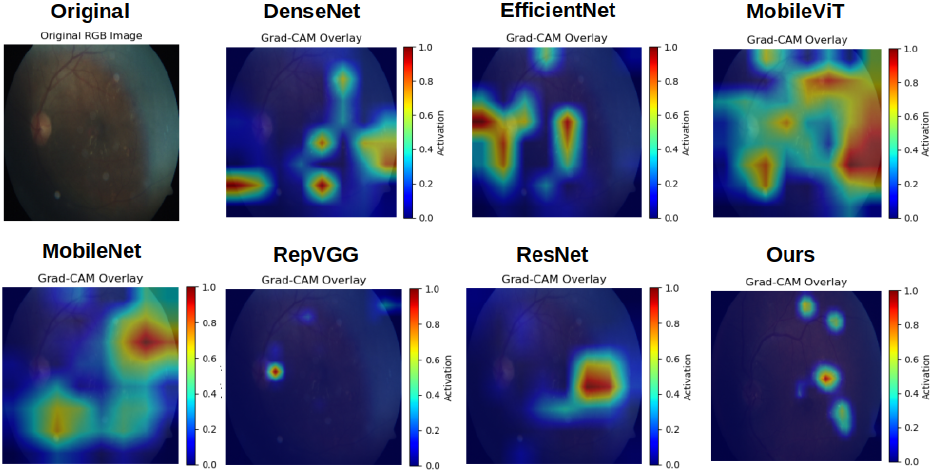
Grad-CAM visual comparisons between our proposed model and other state-of-the-art image classification approaches on the Diabetic Retinopathy dataset.

**Figure 5:**
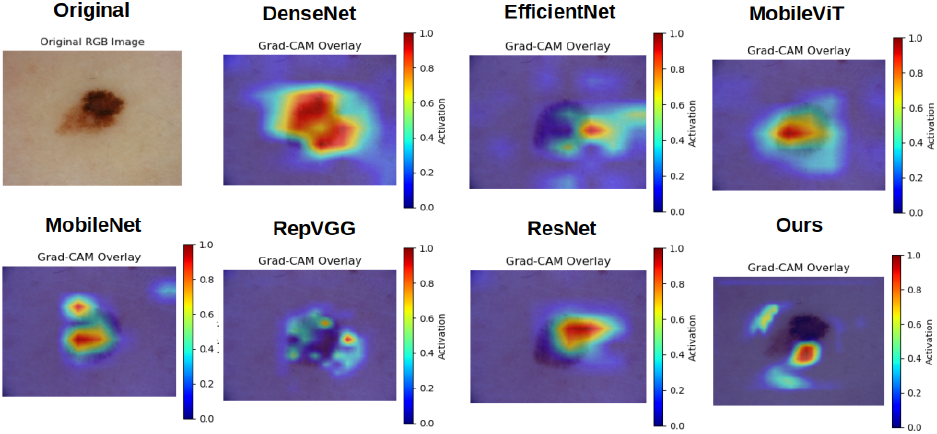
Grad-CAM visual comparisons between our proposed model and other state-of-the-art image classification approaches on the Skin Cancer dataset.

These qualitative results demonstrate that the attention mechanisms integrated into the network, coupled with Grad-CAM analysis, provide interpretable and clinically meaningful explanations for predictions. They also help identify cases where the model may rely on spurious features, guiding future refinement of both architecture and training strategies for enhanced reliability.

### Reliability Diagram

Figure 6 illustrates the reliability diagrams for multiple datasets, including Diabetic-Retinopathy, Kvasir, Oocytes, and Skin-Cancer. Each plot compares the model’s predicted confidence (X-axis) with the corresponding empirical accuracy (Y-axis), providing insights into calibration quality. Ideally, a well-calibrated model’s reliability curve aligns closely with the diagonal reference line, indicating that predicted probabilities accurately reflect true likelihoods. The diagrams show that, across most datasets, the model’s predictions exhibit reasonable calibration, though slight deviations from the diagonal suggest instances of overconfidence or underconfidence in certain confidence intervals.

**Figure 6:**
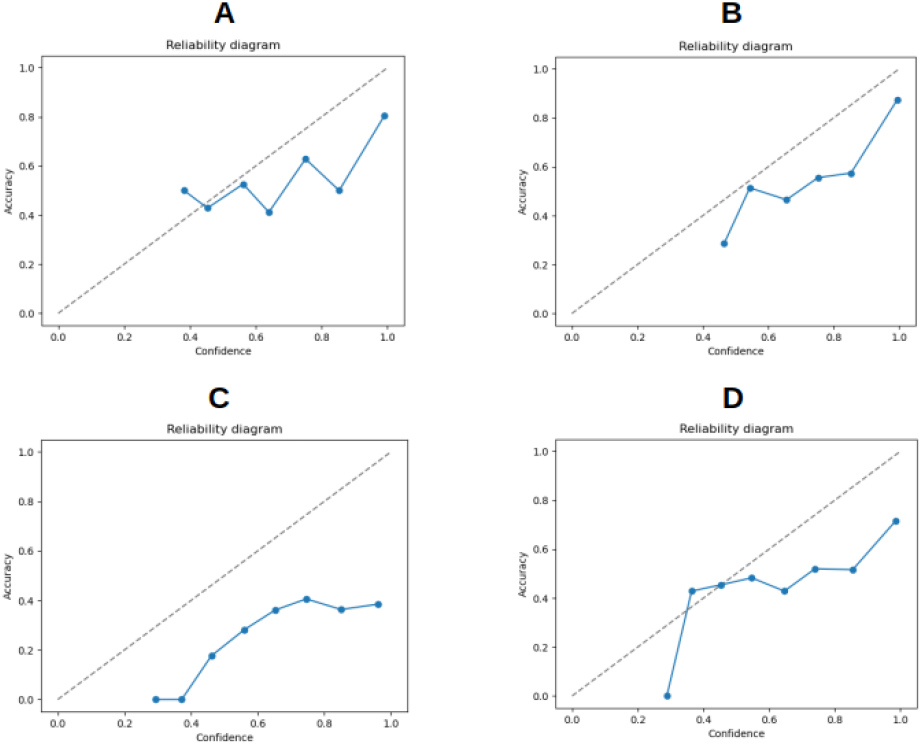
Reliability Diagram using datasets - A: Diabetic-Retinopathy, B: Kvasir, C: Oocytes, and D: Skin-Cancer. The Y-axis represents accuracy, and the X-axis represents confidence.

### Top Uncertain and Certain Samples

To gain deeper insights into the reliability of the proposed model, we further examine its predictive uncertainty on the Diabetic-Retinopathy dataset. Figure 7 presents the five most uncertain and the five most certain cases as estimated by our framework. The most certain cases exhibit high-confidence predictions consistent with the ground-truth labels, underscoring the model’s robustness in recognizing clear diagnostic patterns. In contrast, the most uncertain cases involve challenging samples—such as those with ambiguous lesion boundaries or visually similar inter-class features—where the model demonstrates lower confidence. This analysis highlights not only the value of incorporating uncertainty estimation in medical image classification but also its potential as a diagnostic tool for flagging cases that may warrant additional expert assessment.

**Figure 7:**
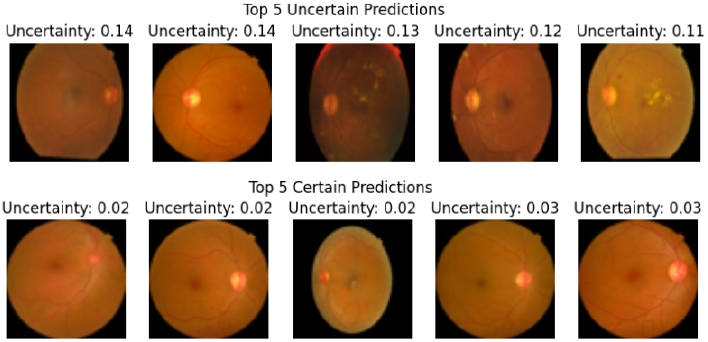
Top row shows the 5 most uncertain samples, while the bottom row shows the 5 most certain samples using the diabetic-retinopathy dataset.

### Ablation Study

Table 2 presents an extensive ablation study conducted on two medical datasets—Diabetic-Retinopathy (A) and Skin-Cancer (B)—to evaluate the impact of different architectural components on model performance. Across both datasets, the proposed model (“Ours”) consistently achieves superior or highly competitive results in nearly all key metrics, including AUC, Accuracy, F1 Score, and MCC, while also demonstrating the lowest computational overhead in terms of inference time and the highest FPS. These results highlight the synergistic benefit of integrating Swirl Attention, Feedback Attention, and Bayesian fully connected (BayesFC) mechanisms, yielding improved predictive reliability, computational efficiency, and uncertainty calibration across diverse medical imaging tasks.

**Table 1:**
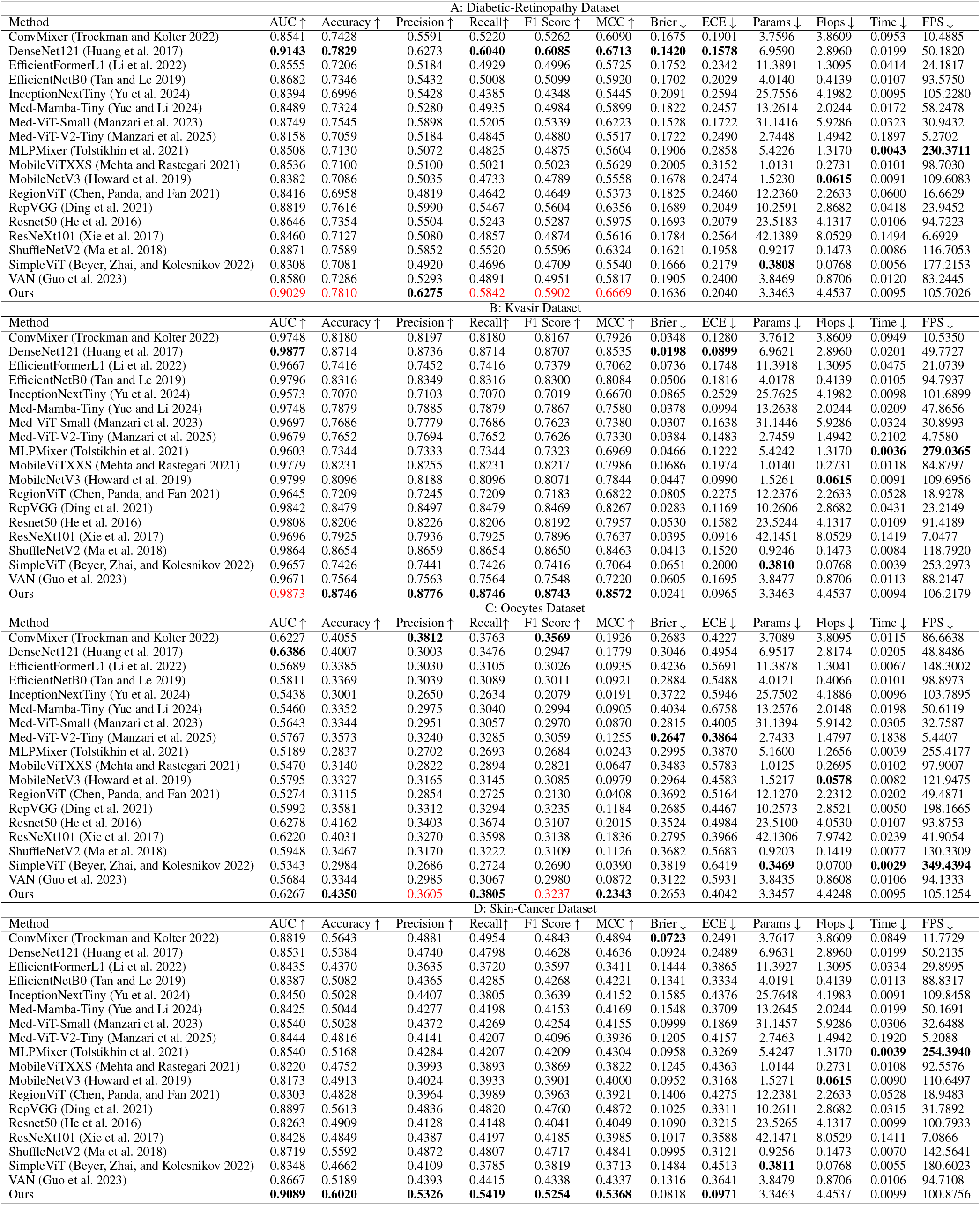
A: Quantitative comparison on the Diabetic Retinopathy dataset. B: Quantitative comparison on the Kvasir dataset. C: Quantitative comparison on the Oocytes dataset. D: Quantitative comparison on the Skin Cancer dataset. The best values are highlighted in bold, while in cases where our model performs the second best, we highlight with the color red.

**Table 2:**
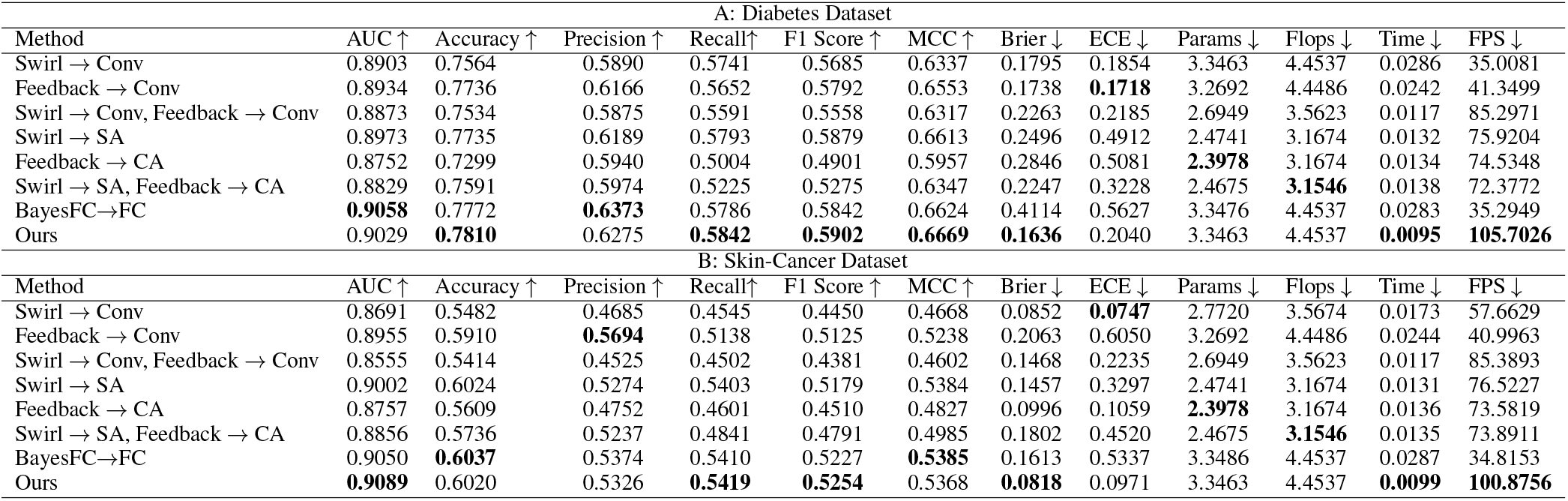
Ablation study using different components in the network architecture. A: Diabetic-Retinopathy dataset. B: Skin-Cancer dataset. Here, Conv, SA, CA, and FC denotes Convolutional layer, Spatial Attention, Channel Attention, and Fully-Connected layer, respectively. The best values are highlighted in bold.

Table 3 presents the ablation analysis comparing the proposed loss function against the conventional Cross-Entropy (CE) loss on the Diabetic-Retinopathy and Skin-Cancer datasets. The results clearly demonstrate the effectiveness of the proposed loss formulation in improving both predictive accuracy and calibration quality. These consistent improvements across datasets highlight the robustness and calibration benefits of the proposed loss, effectively balancing classification accuracy and uncertainty awareness in medical diagnosis tasks.

**Table 3:**
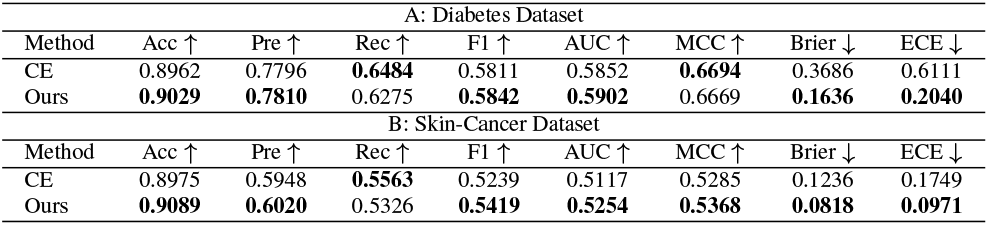
Ablation study using loss function. A: Diabetic-Retinopathy dataset. B: Skin-Cancer dataset. Here CE denotes the Cross-Entropy loss. The best values are highlighted in bold.

### Failure Cases

Despite the overall strong performance of the proposed uncertainty-aware, attention-augmented framework, certain failure cases were observed across the evaluated datasets. In the Diabetic Retinopathy and Skin-Cancer datasets, intermediate stages or borderline morphological variations were occasionally misclassified due to overlapping feature representations between adjacent classes (e.g., Mild vs. Moderate DR). In dermoscopic images from the Skin Cancer dataset, lesions with atypical pigmentation or small size were occasionally misclassified, highlighting the challenge of rare or underrepresented patterns.

We observe instances of both False Positives and False Negatives in Figure 8. False Positives occur when the model’s predictions are correct, but Grad-CAM reveals that the model is focusing on irrelevant or inaccurate regions of the image. False Negatives, on the other hand, occur when the model’s predictions are incorrect, even though Grad-CAM highlights the correct or semantically relevant regions of the image.

**Figure 8:**
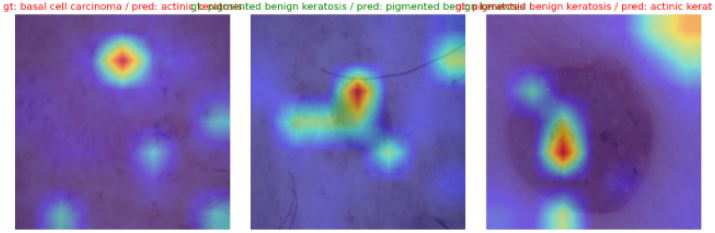
Illustration of some failure cases using GradCAM-based attention maps along with true and predicted labels using the Skin-Cancer dataset.

These failure cases underscore the importance of high-quality imaging, sufficient class representation, and careful preprocessing. They also indicate potential avenues for future work, including multi-modal data integration, improved attention-guided feature extraction, and uncertainty-driven active learning to better handle ambiguous or visually challenging samples.

## Conclusions

In this work, we propose an uncertainty-aware, attention-augmented neural network for medical image classification that combines multi-scale SwirlAttention and FeedBack-Attention modules with a Bayes-by-Backprop probabilistic classifier. The framework enables both discriminative feature learning and reliable uncertainty estimation, addressing the overconfidence of conventional deterministic models. Extensive experiments on four diverse datasets—including Diabetic Retinopathy, Kvasir, Skin Cancer, and fused multifocal Oocyte images—demonstrate improved predictive accuracy, calibration, and interpretability compared to state-of-the-art CNN and transformer-based models. The results highlight the potential of integrating attention mechanisms with Bayesian inference for robust and clinically reliable medical image analysis.

## Data Availability

All data produced are available online at

https://datasets.simula.no/kvasir/

https://www.kaggle.com/datasets/sovitrath/diabetic-retinopathy-224x224-2019-data

https://www.kaggle.com/datasets/nodoubttome/skin-cancer9-classesisic

